# Long-Term Safety and Efficacy of a Highly Purified Plant-Based Nutraceutical For Improving Clinical Parameters of Liver Function in Healthy Participants: A Randomized, Double-Blind, Placebo-Controlled Clinical Trial

**DOI:** 10.1101/2025.09.26.25336688

**Authors:** Ghanashyam Patel, Saumin Shah, Christopher R. D’Adamo

## Abstract

**Background:** Liver function is foundational to human health. While numerous herbal extracts have individually been shown to improve liver function, there are relatively few studies on combinations of herbal ingredients that may have synergistic effects. In addition, capsule fatigue is a common problem when aiming to incorporate multiple ingredients to support liver health. With these gaps in mind, the goal of this study was to evaluate the safety and efficacy of a highly purified plant-based nutraceutical containing organic herbal extracts of turmeric, dandelion, milk thistle, and ginger on liver function parameters.

**Methods:** A randomized, placebo-controlled clinical trial was conducted to evaluate the efficacy of this nutraceutical liquid formula. Healthy adults were randomized 1:1 to receive the nutraceutical product or placebo (both 60 mL) twice daily for 180 days. The primary endpoint was change in liver function tests (alanine aminotransferase [ALT], aspartate aminotransferase [AST], alanine phosphatase [ALP], gamma-glutamyl transferase [GGT]) from baseline to end of study. Safety was also evaluated. The primary endpoint was compared between study arms via analysis of covariance.

**Results:** In total, 130 participants enrolled in the trial; 65 were randomized to each group. Mean improvements in ALT, AST, ALP, and GGT were significantly greater with the nutraceutical product than placebo (P≤0.001). No adverse events were reported.

**Discussion:** Supplementation with this liquid nutraceutical formula improved the liver enzyme profile of healthy individuals, suggesting that it may mitigate a trend toward higher liver enzyme levels over time in a convenient manner that fosters compliance.

**Clinical trial registration:** Clinical Trial Registry of India (https://ctri.nic.in/Clinicaltrials/login.php; CTRI/2023/06/068839)

## Introduction

The liver has many functions that are important to human health, including detoxifying the blood, processing nutrients from food, regulating blood sugar levels and bile production, storing important vitamins, producing proteins involved in blood clotting, helping to maintain a healthy balance of cholesterol in the blood, and producing immune cells [1].

Liver health is typically assessed by measuring serum liver enzymes such as alanine aminotransferase (ALT) and aspartate aminotransferase (AST), as well as markers of cholestasis (alkaline phosphatase and gamma-glutamyl transferase) and other markers. The overall prevalence of elevated liver enzymes has been increasing and is estimated to be approximately 10% of the US population overall [2], with a markedly higher among prevalence in certain subgroups [3–5]. The prevalence of elevated liver enzymes among overweight and obese individuals, in particular, may be as high as 50%. In light of this increasing prevalence, maintenance of liver health has become an especially an important component of overall well-being. Healthy lifestyle choices, such as eating healthy foods, maintaining a healthy weight, incorporation of physical activity, and avoidance of smoking and alcohol are key to support and enhance liver health [6–10].

In addition to these general lifestyle practices, dietary and herbal supplements have become increasingly popular as a means to support liver health. There are numerous herbal extracts that have demonstrated benefits for liver health. For example, turmeric (*Curcuma longa*) has been shown to promote healthy liver cell metabolism and to support tissues, including liver, from oxidative stress [11]. Dandelion (*Taraxacum officinale*) has been shown to help facilitate the metabolism of lipids and insulin [12]. Ginger (*Zingiber officinale*) has been found to support healthy liver function, and to have antioxidant properties [13, 14]. Milk thistle (*Silybum marianum*) has long been used to support the liver’s filtration process and to promote overall liver health [15]. Studies on medication adherence frequently report that polypharmacy, or the need to take multiple pills each day, and pill size/difficulty in swallowing negatively affect treatment adherence [16, 17]. This “capsule fatigue” may also apply to adherence to herbal supplement regimens.

Accordingly, the goal of this study was to evaluate the safety and effectiveness of a portable, nutraceutical liquid supplement formulation containing a mix of herbal extracts previously shown to benefit liver function. The research team hypothesized that the convenience and ease of use this novel delivery vehicle will improve compliance, as it eliminates the need for consumption of multiple individual herbal supplements in capsule form, thereby reducing capsule fatigue. Further, the nutraceutical product is comprised of organic ingredients. Conventionally grown herbal supplements often contain pesticide residues [18], which may be a health risk for consumers, given that chronic pesticide exposure and consumption may elevate the risk of a variety of negative health outcomes [19–21]. The research team hypothesized that the nutraceutical liquid formulation would be safe and improve a range of measures of liver function under study. To this end, the study aimed to evaluate the effects of repeated supplementation of the nutraceutical liquid formulation on liver function in healthy individuals.

## 2. Patients and Methods

### 2.1. Patients

Healthy participants aged between 18 and 60 years with normal liver function (i.e., liver function test values were within the upper limit of normal) were eligible for study enrollment. The main exclusion criteria were a known or suspected hypersensitivity or intolerance to herbal products (e.g., turmeric, dandelion, ginger, or milk thistle extract); a liver-related disease or abnormality; a history of regular alcohol consumption (>14 drinks/week for females; >21 drinks/week for males), alcoholism, or drug abuse; and use of drugs or medications known to affect hepatic function in the 4 weeks prior to randomization.

The clinical trial protocol was approved by the Institutional Ethics Committee at the Unity Trauma Centre and Intensive Care Unit at Unity Hospital, Gujarat, India (ECR/1226/Inst/GJ/2019, approved on 22 June 2023). The study was conducted according to the principles outlined in the Declaration of Helsinki and the International Council for Harmonisation Good Clinical Practice guidelines, along with the local regulatory requirements of Good Clinical Practice for Clinical Research in India, the New Drugs and Clinical Trial Rules, the Indian Council of Medical Research National Ethical Guidelines for Biomedical and Health Research Involving Human Participants, and relevant guidelines of Ayurveda, Yoga and Naturopathy, Unani, Siddha and Homeopathy (AYUSH) Good Clinical Practice and the Clinical Evaluation Ayurvedic Interventions by AYUSH. All study participants provided written informed consent. The study was registered at the Clinical Trial Registry of India (CTRI/2023/06/068839) on June 27, 2023. There was no patient or public involvement in the design, conduct, or reporting of the trial.

### 2.2. Study design and treatments

This was a prospective, randomized, double-blind, placebo-controlled, parallel group, exploratory clinical study to evaluate the effectiveness and safety of Dose for your Liver^®^ (Eetho Brands Inc., Marina Del Rey, CA, USA), a commercially available dietary supplement. This study was conducted at a single center (Unity Trauma Centre and Intensive Care Unit at Unity Hospital, Gujarat, India) between July 2023 and March 2024. Participants underwent a 7-day screening phase during which baseline clinical data and laboratory values were collected.

Participants were then randomized 1:1 (Day 1) to receive either the test product 60 mL twice daily or placebo 60 mL twice daily for 180 days. Participants received a 30-day supply of the study supplement and a subject diary card. Follow-up visits were conducted every 30 days (Days 31, 61, 91, 121, 151, 181) where the participant received an additional 30-day supply of the study treatment and a new subject diary card. Follow-up visits included a physical examination, assessment of subject diary card and study treatment compliance, and safety monitoring. Effectiveness assessments included liver function tests (alanine aminotransferase [ALT], aspartate aminotransferase [AST], alkaline phosphatase [ALP], and gamma-glutamyl transferase [GGT]), AST to platelet ratio index (APRI) score, complete blood count (CBC) parameters (hemoglobin, red blood cells, white blood cells, and platelet count), lipid profile (total cholesterol, high density lipoproteins [HDL], low density lipoproteins [LDL], very low density lipoproteins [VLDL]), kidney function tests (serum creatinine and blood urea nitrogen [BUN]), inflammatory markers (IgA, IgA, C-reactive protein [CRP]), and fasting blood sugar (FBS), C-peptide, and ferritin levels. Vital signs, laboratory tests, and adverse events (AEs) and serious AEs (SAEs) were monitored for safety. AEs and SAEs were coded and classified using the Medical Dictionary for Regulatory Activities, version 24.1.

The test product (Dose for your Liver^®^; Eetho Brands Inc., Marina Del Rey, CA, USA) is a highly purified plant-based nutraceutical designed to support liver function. It contains organic turmeric extract, organic dandelion powder, organic milk thistle extract, and organic ginger powder. The placebo product contained reverse osmosis water, erythritol, citric acid, monk fruit extract, and natural orange flavoring. The study intervention and placebo were identical in appearance, taste, and packaging. Each serving of the test product or placebo was provided as a 60 mL oral solution and was to be consumed twice daily (morning and evening) with a meal for the 180-day treatment period. A 30-day supply was distributed at each study/follow-up visit.

The randomization sequence was generated using the Experimental Design Generator and Randomizer (John Innes Centre, Norwich, England). Both the participants and the investigators were blinded to the intervention assignments per the double-blind study design. This process aimed to eliminate potential biases associated with participants’ expectations and investigators’ assessments, thereby enhancing the internal validity of the study. The biostatistician, independent of this study, securely maintained the blinding of randomization codes, ensuring the concealment of group assignments until the completion of the data analysis phase.

The following medications were prohibited during the study: statins, fibrates, bile acid sequestrants, niacin (nicotinic acid) and derivatives, ezetimibe, alirocumab, and evolocumab.

### 2.3. Study endpoints

The primary endpoint was change from baseline (i.e., improvement) in liver function test values (ALT, AST, ALP, GGT) from baseline to end of study. Secondary endpoints included change from baseline to end of study in APRI, lipid profile parameters, complete blood count parameters, kidney function tests, inflammatory markers, and other parameters (FBS, ferritin, C-peptide). The proportion of patients who experienced an improvement in the clinical parameters listed above was also assessed. AEs and SAEs were monitored for safety.

### 2.4. Statistical analysis

Considering the exploratory nature of this study, no formal sample size calculation was performed. It was thought that 120 participants (60 in each group) would be sufficient to evaluate the effectiveness and safety of the test product compared with placebo.

Efficacy analyses were conducted using the per protocol population, which included all randomized participants who were administered ≥80% of the planned test product or placebo doses through the end of the study, who had no protocol deviation(s) that would affect treatment evaluation, and completed all the designated study visits. The safety population included all randomized participants who received at least one dose of the test product or placebo post-randomization.

Descriptive statistics were used to summarize continuous data, and categorical variables were summarized using frequency and percent. Chi square test (categorical variables) and Welch’s t-test (continuous variables) were performed to evaluate differences in participant characteristics at baseline. Chi-square tests were performed to assess the proportion of patients who had an improvement in their ALT, AST, ALP, GGT, APRI, total cholesterol, triglycerides, LDL, VLDL, CRP, IgA, IgE, and ferritin values from baseline to end of study. For the chi-square tests and Welch’s t-test, a p-value of <0.05 was considered statistically significant. Analysis of covariance (ANCOVA) was used to adjust for baseline values of the primary and secondary outcomes in the comparison of treatment effects between groups. Since the primary outcome of liver enzymes consisted of four analytes (ALT, AST, ALP, GGT), a Bonferroni correction was applied to control the family-wise error rate. Thus, a p-value of <0.0125 was considered statistically significant. Statistical analyses were performed using SAS version 9.4 (SAS Institute; Cary, NC, USA).

## 3. Results

### 3.1 Participants

The first participant was enrolled on July 5, 2023, and the study was completed on June 24, 2024. A total of 140 participants were screened, 10 participants withdrew their consent prior to randomization, and 130 participants were randomized and completed the study (test product, n = 65; placebo, n = 65) (**Fig. 1**). All 130 participants attended all the scheduled study visits and were included in the effectiveness (per protocol population) and safety (safety population) analyses. In the test product and placebo groups, the respective mean ± standard deviation (SD) age was 36.5 ± 11.6 years and 36.8 ± 10.7 years, the mean ± SD Body Mass Index was 23.7 ± 2.8 mg/kg^2^ and 23.4 ± 2.6 mg/kg^2^, and 69.2% (n = 45) and 55.4% (n = 36) of participants were male (**Table 1**). While the baseline ALT and AST levels were within the normal range (7-56 IU/L and 10-40 IU/L, respectively) they were near the upper limit. All patients had a treatment compliance rate between 80% and 100%. The lowest and highest compliance rates for the study were 81.1% and 98.9%, respectively.

**Figure 1.**
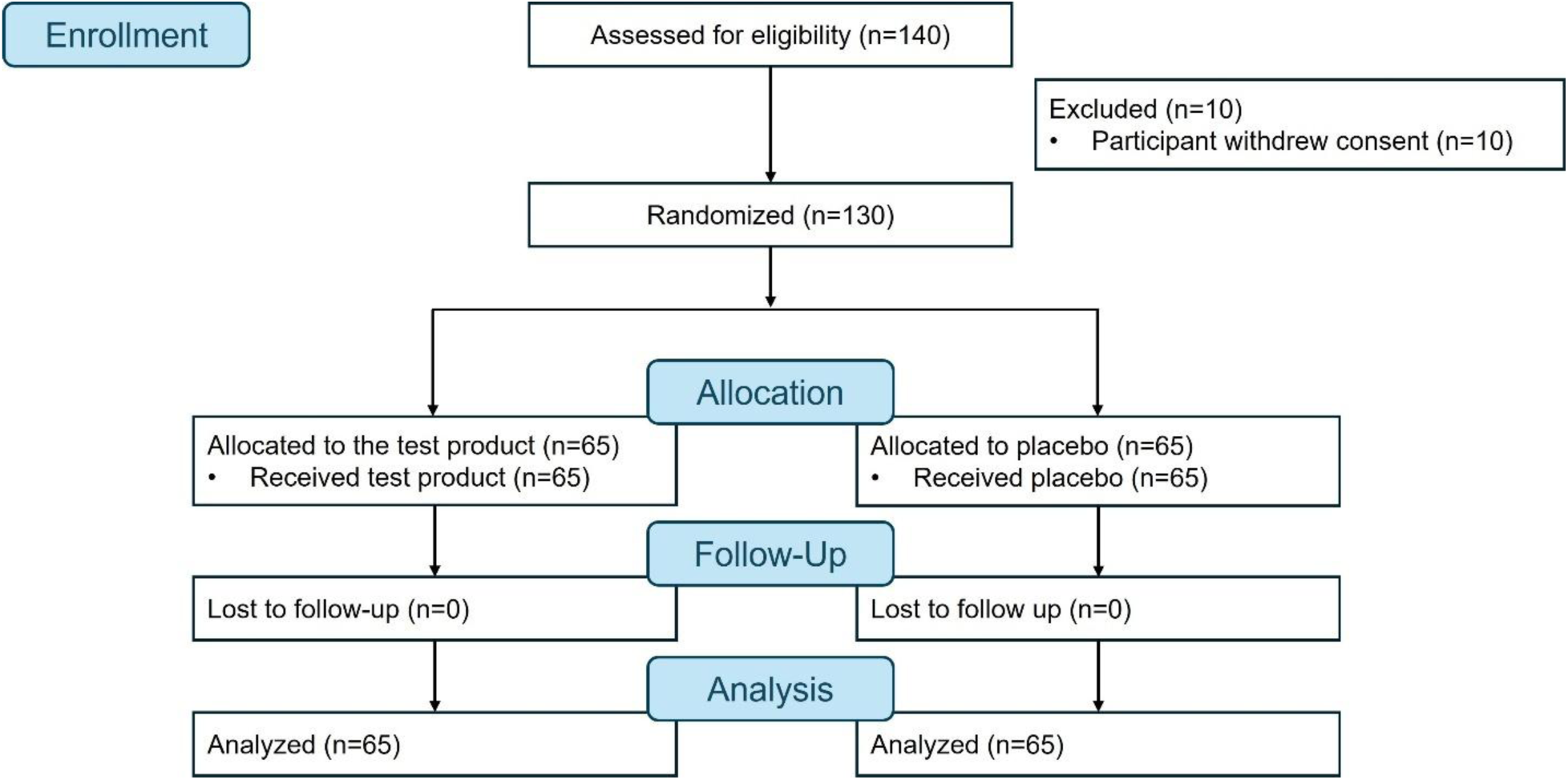
Flow diagram.

**Table 1.**
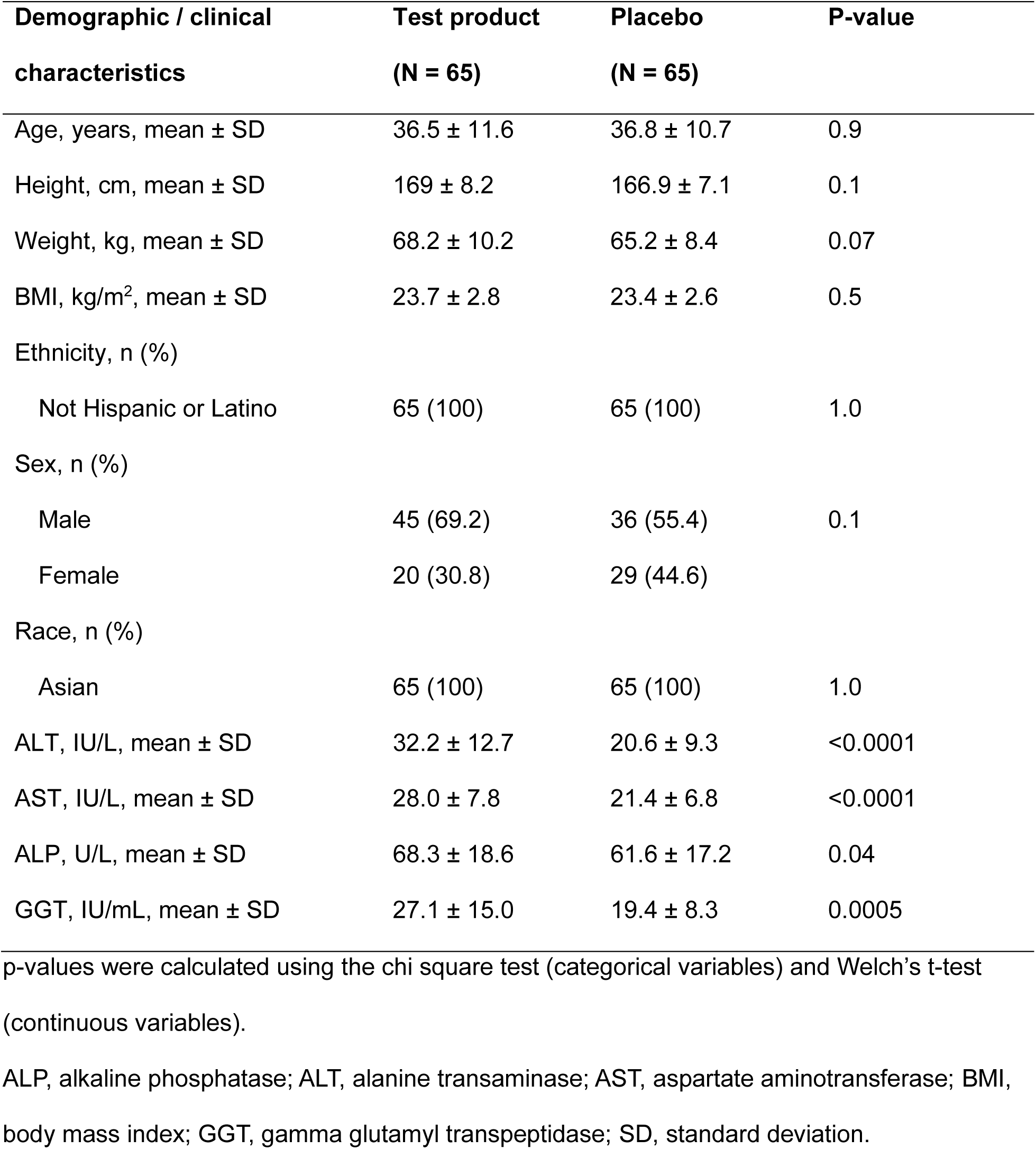
Participant demographic and clinical characteristics.

| Demographic / clinical characteristics | Test product<br>(N = 65) | Placebo<br>(N = 65) | P-value |
| --- | --- | --- | --- |
| Age, years, mean ± SD | 36.5 ± 11.6 | 36.8 ± 10.7 | 0.9 |
| Height, cm, mean ± SD | 169 ± 8.2 | 166.9 ± 7.1 | 0.1 |
| Weight, kg, mean ± SD | 68.2 ± 10.2 | 65.2 ± 8.4 | 0.07 |
| BMI, kg/m<sup>2</sup>, mean ± SD | 23.7 ± 2.8 | 23.4 ± 2.6 | 0.5 |
| Ethnicity, n (%) |  |  |  |
| Not Hispanic or Latino | 65 (100) | 65 (100) | 1.0 |
| Sex, n (%) |  |  |  |
| Male | 45 (69.2) | 36 (55.4) | 0.1 |
| Female | 20 (30.8) | 29 (44.6) |  |
| Race, n (%) |  |  |  |
| Asian | 65 (100) | 65 (100) | 1.0 |
| ALT, IU/L, mean ± SD | 32.2 ± 12.7 | 20.6 ± 9.3 | <0.0001 |
| AST, IU/L, mean ± SD | 28.0 ± 7.8 | 21.4 ± 6.8 | <0.0001 |
| ALP, U/L, mean ± SD | 68.3 ± 18.6 | 61.6 ± 17.2 | 0.04 |
| GGT, IU/mL, mean ± SD | 27.1 ± 15.0 | 19.4 ± 8.3 | 0.0005 |

p-values were calculated using the chi square test (categorical variables) and Welch's t-test (continuous variables).
ALP, alkaline phosphatase; ALT, alanine transaminase; AST, aspartate aminotransferase; BMI, body mass index; GGT, gamma glutamyl transpeptidase; SD, standard deviation.

### 3.2 Effectiveness endpoints

For the primary endpoint, change in liver function test values from baseline to end of study was significantly better with the test product compared with placebo (**Table 2**). The mean ± SD change in liver function test values from baseline to end of study for the test product and placebo groups was a respective –6.0 ± 5.2 IU/L (i.e., decrease from baseline to end of study) and +10.6 ± 15.0 IU/L (i.e., increase from baseline to end of study) for ALT (between group difference, p <0.001), –6.5 ± 11.0 IU/L and +4.2 ± 11.7 IU/L for AST (p <0.001), +2.3 ± 28.5 U/L and +15.6 ± 30.5 U/L for ALP (p = 0.01), and –4.0 ± 19.8 IU/mL and +9.8 ± 19.6 IU/mL for GGT (p <0.001). Overall, a numerically higher proportion of participants in the test product group versus placebo group had an improvement in their liver enzyme values (baseline to end of study), and the difference was significant for ALT, AST, and GGT, but not ALP. Improvement in ALT was observed for 61.5% (40/65) of patients in the test product group and 13.8% (9/65) of patients in the placebo group (chi-square p<0.001), improvements in AST were observed for 76.9% (50/65) and 21.5% (14/65) (chi-square p<0.001), improvements in ALP were observed for 44.6% (29/65) and 30.8% (20/65) (chi-square 0.15), and improvements in GGT were observed for 60.0% (39/65) and 24.6% (16/65) (chi-square p<0.001) (**Fig. 2**).

**Figure 2.**
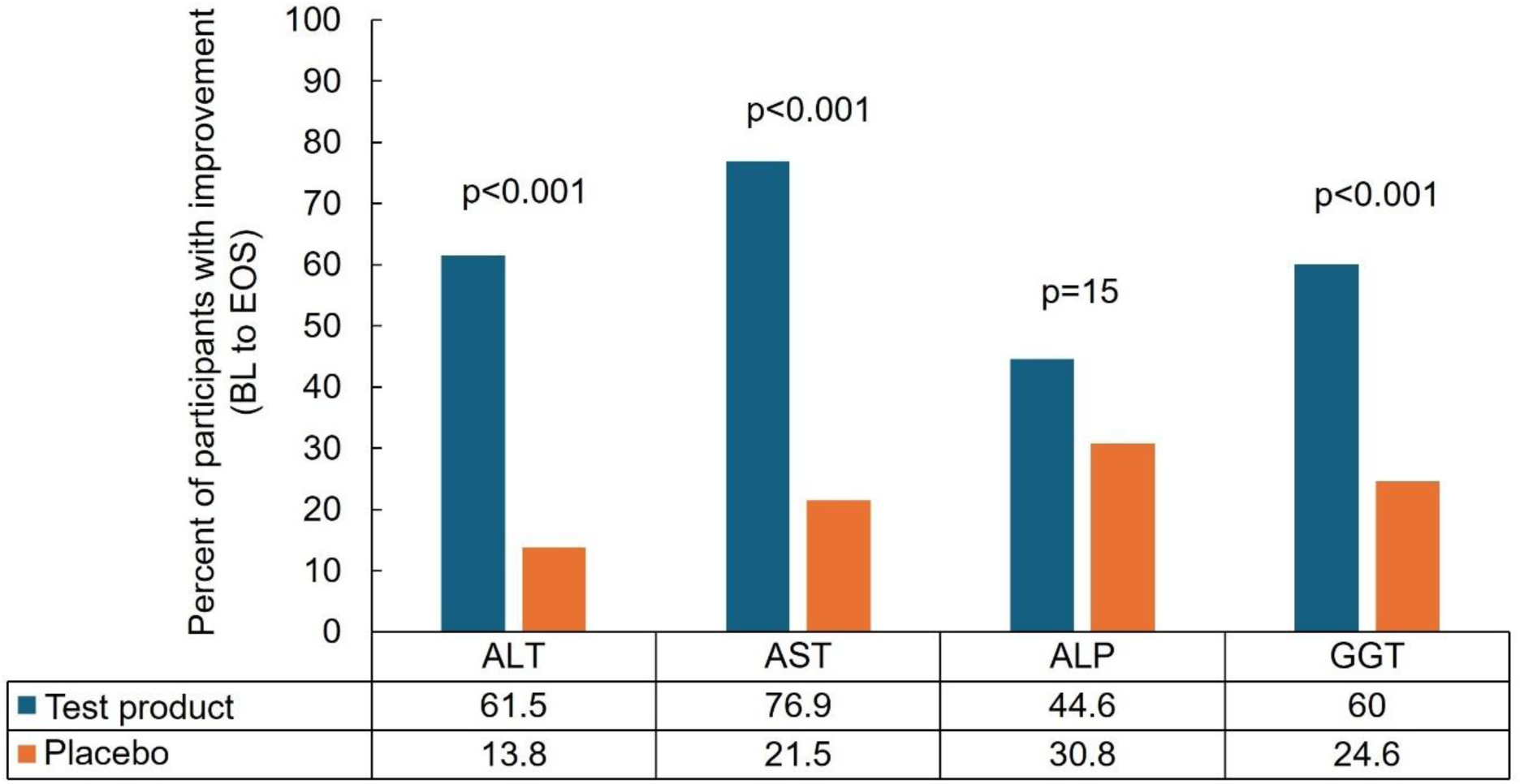
Percent of participants with improvement from baseline to end of study in liver function tests. p-values were calculated using the chi-square test. ALP, alkaline phosphatase; ALT, alanine transaminase; AST, aspartate aminotransferase; BL, baseline; EOS, end of study; GGT, gamma-glutamyl transferase.

**Table 2.** Mean change from baseline to end of study in liver function test parameters (primary endpoint).

| Liver function test<br>parameter | Test product<br>(N = 65) | Placebo<br>(N = 65) | P-value |
| --- | --- | --- | --- |
| ALT, IU/L | -6.0 ± 5.2 | +10.6 ± 15.0 | <0.001 |
| AST, IU/L | -6.5 ± 11.0 | +4.2 ± 11.7 | <0.001 |
| ALP, U/L | +2.3 ± 28.5 | +15.6 ± 30.5 | 0.01 |
| GGT, IU/mL | -4.0 ± 19.8 | +9.8 ± 19.6 | <0.001 |

Data are shown as mean ± SD. Data are shown as mean ± SD. P-values were calculated using the Analysis of Covariance. ALP, alkaline phosphatase; ALT, alanine transaminase; AST, aspartate aminotransferase; GGT, gamma glutamyl transpeptidase; SD, standard deviation.

Mean ± SD values for change from baseline to end of study for the secondary endpoints are shown in **Table 3**. Improvement in APRI score, total cholesterol, LDL, CRP, IgA, IgE, and ferritin levels from baseline to end of study was significantly greater among participants in the test product group compared with the placebo group (p <0.001 for all). A numerically but not significantly higher proportion of participants in the test product group versus the placebo group had an improvement in these values APRI score (66.2% vs 38.5%; chi-square p=0.22), total cholesterol (67.7% vs 56.9%; chi-square p=0.66), LDL (60.0% vs 33.9%; chi-square p=0.15), IgA (81.5% vs 69.2%; chi-square p=0.69), IgE (78.5% vs 52.3%; chi-square p=0.73), CRP (46.2% vs 26.2%; chi-square p=0.42), and ferritin (72.3% vs 58.5%; chi-square p=0.63) values from baseline to end of study (**Fig. 3**). Improvement in all other secondary endpoint parameters was not statistically significant between the two groups (**Table 3**).

**Figure 3.**
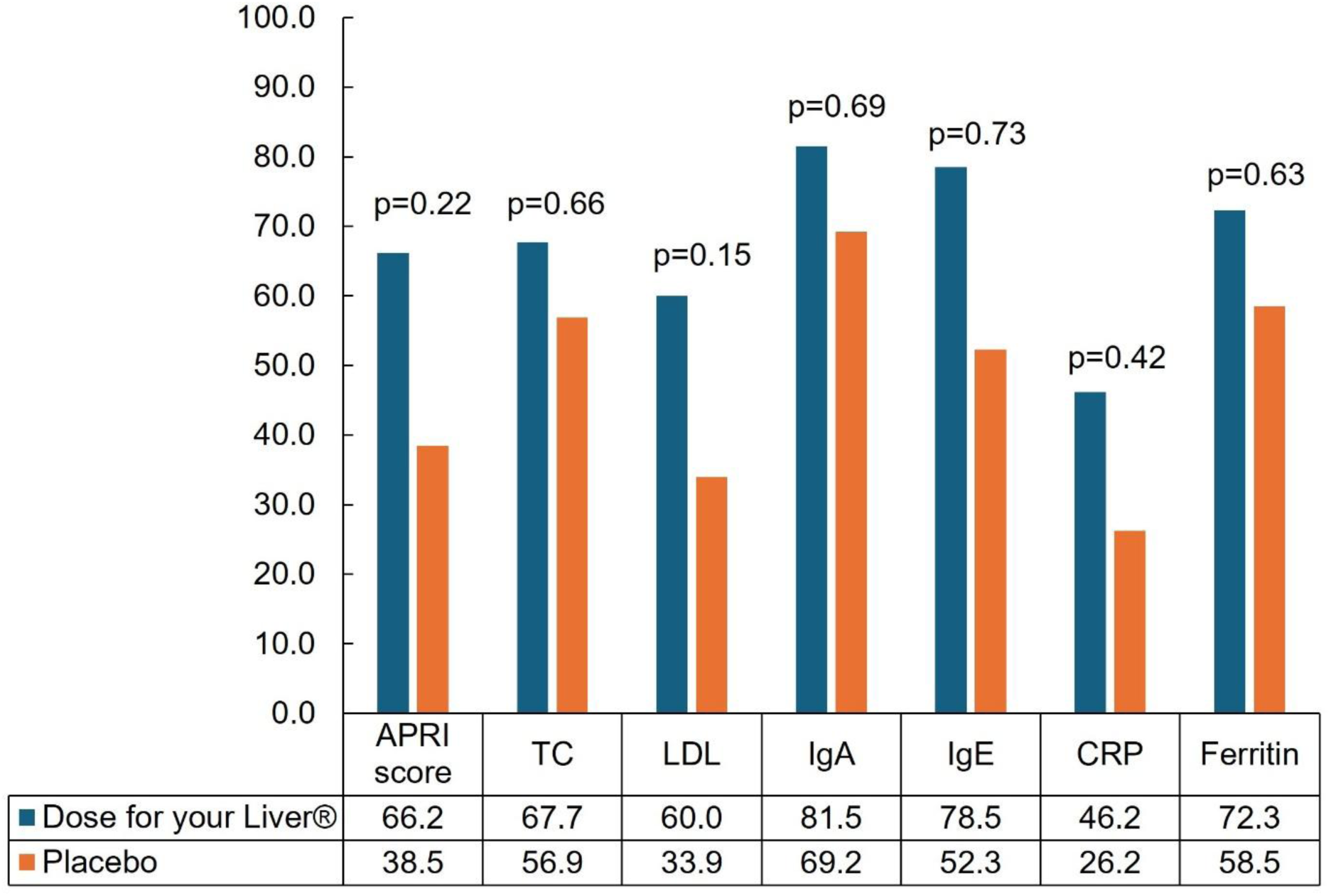
Percent of participants with significant improvement from baseline to end of study in the secondary efficacy endpoint parameters of APRI, lipid measures (TC, LDL), immune measures (IgA, IgE, CRP), and ferritin. p-values were calculated using the chi-square test. APRI, aspartate aminotransferase to platelet ratio index; BL, baseline; CRP, C-reactive protein; EOS, end of study; Ig, immunoglobin; LDL, low-density lipoproteins; TC, total cholesterol.

**Table 3.**
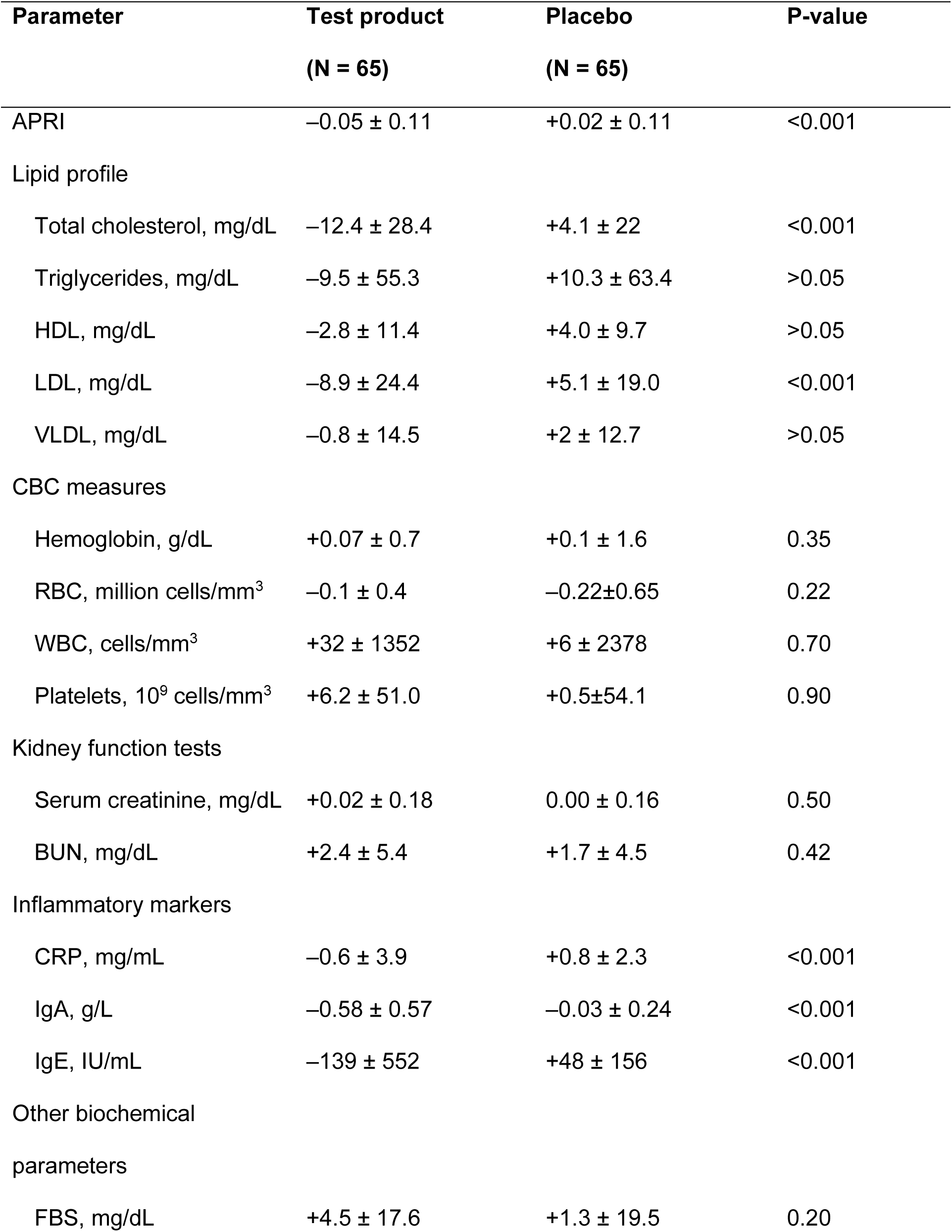
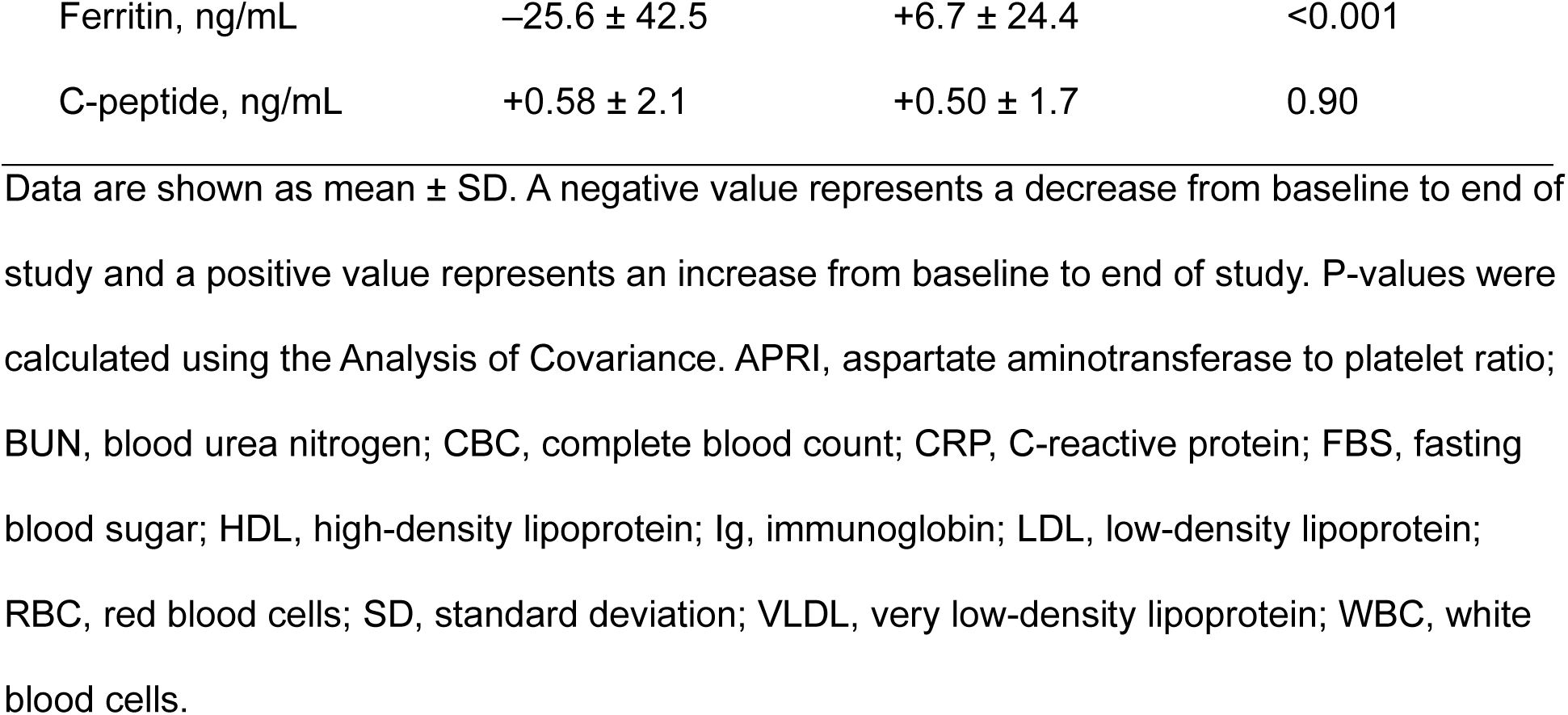
Mean change from baseline to end of study in secondary endpoint parameters.

| Parameter | Test product<br>(N = 65) | Placebo<br>(N = 65) | P-value |
| --- | --- | --- | --- |
| APRI | -0.05 ± 0.11 | +0.02 ± 0.11 | <0.001 |
| Lipid profile |  |  |  |
| Total cholesterol, mg/dL | -12.4 ± 28.4 | +4.1 ± 22 | <0.001 |
| Triglycerides, mg/dL | -9.5 ± 55.3 | +10.3 ± 63.4 | >0.05 |
| HDL, mg/dL | -2.8 ± 11.4 | +4.0 ± 9.7 | >0.05 |
| LDL, mg/dL | -8.9 ± 24.4 | +5.1 ± 19.0 | <0.001 |
| VLDL, mg/dL | -0.8 ± 14.5 | +2 ± 12.7 | >0.05 |
| CBC measures |  |  |  |
| Hemoglobin, g/dL | +0.07 ± 0.7 | +0.1 ± 1.6 | 0.35 |
| RBC, million cells/mm<sup>3</sup> | -0.1 ± 0.4 | -0.22±0.65 | 0.22 |
| WBC, cells/mm<sup>3</sup> | +32 ± 1352 | +6 ± 2378 | 0.70 |
| Platelets, 10<sup>9</sup> cells/mm<sup>3</sup> | +6.2 ± 51.0 | +0.5±54.1 | 0.90 |
| Kidney function tests |  |  |  |
| Serum creatinine, mg/dL | +0.02 ± 0.18 | 0.00 ± 0.16 | 0.50 |
| BUN, mg/dL | +2.4 ± 5.4 | +1.7 ± 4.5 | 0.42 |
| Inflammatory markers |  |  |  |
| CRP, mg/mL | -0.6 ± 3.9 | +0.8 ± 2.3 | <0.001 |
| IgA, g/L | -0.58 ± 0.57 | -0.03 ± 0.24 | <0.001 |
| IgE, IU/mL | -139 ± 552 | +48 ± 156 | <0.001 |
| Other biochemical parameters |  |  |  |
| FBS, mg/dL | +4.5 ± 17.6 | +1.3 ± 19.5 | 0.20 |
| Ferritin, ng/mL | -25.6 ± 42.5 | +6.7 ± 24.4 | <0.001 |
| C-peptide, ng/mL | +0.58 ± 2.1 | +0.50 ± 1.7 | 0.90 |

Data are shown as mean ± SD. A negative value represents a decrease from baseline to end of study and a positive value represents an increase from baseline to end of study. P-values were calculated using the Analysis of Covariance. APRI, aspartate aminotransferase to platelet ratio; BUN, blood urea nitrogen; CBC, complete blood count; CRP, C-reactive protein; FBS, fasting blood sugar; HDL, high-density lipoprotein; Ig, immunoglobulin; LDL, low-density lipoprotein; RBC, red blood cells; SD, standard deviation; VLDL, very low-density lipoprotein; WBC, white blood cells.

### 3.3. Safety

There were no AEs, SAEs, or deaths reported in either study group. Additionally, there were no clinically significant findings for vital signs or laboratory parameters, or during physical examination for either study group.

## 4. Discussion

While previous reports have indicated the potential benefits of turmeric, dandelion, milk thistle, and ginger on the liver individually, to the authors’ knowledge, this is the first report of the effects of these extracts combined in any form. Moreover, this is the first report of these extracts combined in a liquid, single-use format. This study demonstrated the effectiveness and safety of the test product. In healthy participants, twice daily supplementation with the test product over 180 days improved liver health from baseline to end of study. This was evidenced by improvements (i.e., greater change from baseline to end of study) in the key liver function tests, ALT, AST, ALP, and GGT, as well as the APRI score, all of which achieved a significantly greater reduction over time in the test product group than in the placebo group. Of note, levels of ALT, AST, ALP, and GGT in the placebo group had numerical increases from baseline to end of study, while these levels generally had numerical decreases with test product supplementation. The test product also improved the lipid profile and levels of inflammatory markers and ferritin. Further, there was no negative effect on hematological (CBC) or kidney function indicators. Similarly, the test product had no significant effect on FBS or C-peptide levels. The safety of repeated supplementation was demonstrated, as there were no AEs or SAEs reported for either study group.

Intragroup comparisons demonstrated that 4.5 times and 3.6 times more participants in the test product than placebo group experienced improvement in ALT and AST levels, respectively. Improvement in these liver-related parameters is likely attributable to the hepatoprotective activity of curcumin, the major bioactive present in the turmeric rhizome. Curcumin may improve hepatic steatosis and block the progression of fatty liver disease through the inhibition of fatty acid synthesis and the biosynthesis of unsaturated fatty acids [22]. The suggested liver-protective effects of the test product are supported by previous studies that have demonstrated a decrease in metabolic biomarkers with curcumin, indicating a potential role in countering NAFLD [23]. The reported antioxidant and anti-inflammatory effects of curcumin may also contribute to lowering ALT and AST levels, especially when given for a longer duration and at higher doses (≥1000 mg/day) [22, 24–26]. Other ingredients in the test product may also be contributing to the improvement in liver enzyme levels. Silymarin, a mixture of flavonolignans extracted from milk thistle, has demonstrated anti-hepatotoxic effects in both animal and human studies [27–29]. It has been reported to reduce mortality among patients with cirrhosis of the liver [27] and to decrease liver enzymes in patients with alcoholic liver disease and chronic viral hepatitis [28]. Both dandelion and ginger have shown hepatoprotective effects, which are thought to be explained by their ability to reduce oxidative stress [30, 31]. Future studies are needed to understand whether there are synergistic effects when these herbal extracts are consumed in combination.

Although the study participants were considered healthy and their baseline AST and ALT levels were within the normal range, the levels were somewhat elevated, possibly indicating subclinical metabolic dysfunction. Further, mean levels of these enzymes were numerically increased from baseline to end of study in the placebo group, suggesting low-level or subclinical liver stress in the absence of active liver support. Progression to liver disease typically starts with steatosis (i.e., fatty liver), then progresses to elevated liver enzyme levels, fibrosis, and, finally, cirrhosis. Thus, reducing liver enzyme levels very early in the process suggests a proactive approach to slowing the progression of liver dysfunction. Importantly, the current standard of care following the observation of increased AST/ALT levels without confirmation of liver fibrosis is to “watch and wait” and recommend lifestyle changes [32]. The test product may provide an option for people who have no clinical symptoms of liver disease but are experiencing AST/ALT levels that are on the high end of the normal range or who have noted a slow increase in enzyme levels over time, with the potential to manage liver stress/early liver dysfunction and to slow progression towards liver disease.

The test product not only prevented the enzyme elevations observed in the placebo group, it reversed them. The enzyme data suggest a subclinical drift toward liver dysfunction for patients in the placebo group (i.e., increasing levels over time), while the test product group showed directional correction of this drift (i.e., decreasing levels over time), indicating the potential for targeted nutrition as an early intervention strategy. These findings also highlight the utility of liver enzymes as sentinel biomarkers for metabolic stress, and how small lifestyle changes have the potential to meaningfully shift liver function towards a more balanced state.

Change from baseline to end of study between the test product and placebo groups was also significant for the APRI score, total cholesterol, LDL, IgA, IgE, CRP, and ferritin levels. APRI score is a non-invasive marker for assessing the severity of liver fibrosis. A numerically but not significantly greater proportion of participants in the test product group had an improvement in APRI score than participants in the placebo group. This further supports the potential benefit of the test product on liver health. A healthy liver plays a key role in maintaining optimal lipid levels in the blood stream. The proportion of participants who had an improvement in their total cholesterol and LDL levels was numerically but not significantly greater with the test product versus placebo. This suggests a potential benefit to liver function with test product supplementation.

Given that the liver plays a role in the immune response to external pathogens, this study also evaluated the effect of test product supplementation on levels of the immune markers IgA, IgG, and CRP. The proportion of participants who experienced improvement in IgA, IgE, and CRP levels was numerically but not significantly higher in the test product group compared with the placebo group. These findings suggest a potential anti-inflammatory effect for the test product.

Twice daily dosing of the test product supplement over 180 days was safe, with no reported AEs or SAEs. Further, the similar findings between the test product and placebo groups for CBC and kidney function tests, as well as FBS and C-peptide levels support the overall safety of the product.

This study had some limitations that should be considered when interpreting the results. This was a single center study; additional studies would be helpful to expand generalizability to other populations. This study only included healthy participants, so the effectiveness of the test product in patients with liver disease is unknown.

## 5. Conclusions

Supplementation with the test product was effective for improving markers of liver health among healthy participants. Participants in the placebo group experienced elevations in AST, ALT, and GGT from baseline, suggesting a trend of increasing subclinical liver stress in healthy individuals over time in the absence of proactive liver support. The test product demonstrated an exceptional safety profile with repeated dosing, indicating that regular consumption is safe. These findings suggest the potential for lowering liver enzyme levels with repeated supplementation of this highly purified plant-based nutraceutical in healthy individuals who may have subclinical metabolic dysfunction or who are experiencing a trend of increasing liver enzyme levels over time.

## Ethics

The study was conducted according to the guidelines of the Declaration of Helsinki, and approved by the Institutional Ethics Committee of the Unity Trauma Centre and Intensive Care Unit at Unity Hospital, Gujarat, India (ECR/1226/Inst/GJ/2019, approved on 22 June 2023). All study participants provided written informed consent.

## Data availability

Data will be made available on request to the corresponding author.

## Acknowledgments

The authors thank Sarah Bubeck, PhD, for providing medical writing support, which was funded by Eetho Brands Inc. This research was funded by Eetho Brands Inc. who were involved in the design and reporting of the trial, but not in the conduct of the trial or analysis of data.

## CRediT authorship contribution statement

Conceptualization, G.P.; methodology, G.P., C.D.; validation, G.P.; investigation, S.S.; data curation, S.S.; writing—original draft preparation, G.P.; writing—review and editing, G.P., S.S., C.D.; visualization, G.P.; supervision, G.P.; project administration, G.P. All authors have read and agreed to the published version of the manuscript.

## Declaration of competing interest

Ghanashyam Patel is currently an employee at Dose (Eetho Brands Inc.). Saumin Shah has no competing interests to declare. Christopher R. D’Adamo is a consultant for Dose (Eetho Brands Inc.). The funders had a role in the design of the study; the collection, analysis, and interpretation of data; the writing of the manuscript; and the decision to publish the results.

## Abbreviations

ANCOVA: Analysis of covariance
AE: Adverse event
ALP: Alkaline phosphatase
ALT: Alanine transaminase
APRI: Aspartate aminotransferase to platelet ratio index
AST: Aspartate aminotransferase
AYUSH: Ayurveda, Yoga and Naturopathy, Unani, Siddha and Homeopathy
BL: Baseline
BUN: Blood urea nitrogen
CBC: Complete blood count
CRP: C-reactive protein
EOS: End of study
FBS: Fasting blood sugar
GGT: Gamma glutamyl transpeptidase
HDL: High-density lipoprotein
Ig: Immunoglobulin
LDL: Low-density lipoprotein
RBC: Red blood cells
SAE: Serious adverse event
SD: Standard deviation
TC: Total cholesterol
TG: Triglyceride
VLDL: Very low-density lipoprotein
WBC: White blood cells
AST: Aspartate aminotransferase

## Notes

### Clinical Trial

CTRI/2023/06/068839

### Author Declarations

The Institutional Ethics Committee of the Unity Trauma Centre and Intensive Care Unit at Unity Hospital, Gujarat, India gave ethical approval for this work (ECR/1226/Inst/GJ/2019, approved on 22 June 2023).

